# Fifteen-year Outcomes of 1,196 Ozaki Procedures

**DOI:** 10.1101/2023.05.08.23289697

**Authors:** Shigeyuki Ozaki, Yasuhiro Hoshino, Shinya Unai, Serge C. Harb, William C. Frankel, Hiromasa Hayama, Mikio Takatoo, Nagaki Kiyohara, Hiroshi Kataoka, Lars G. Svensson, Jeevanantham Rajeswaran, Eugene H. Blackstone, Gösta B. Pettersson

## Abstract

**Background:** Introduced in 2007, the Ozaki procedure has become an attractive option for aortic valve disease. Our objective was to investigate outcomes of the Ozaki procedure in the original Ozaki cohort.

**Methods:** From April 2007 to May 2021, 1,196 consecutive Ozaki procedures were performed at Toho University Ohashi Medical Center. Patient age ranged from 11 to over 90 years, 484 (60%) were male, 50 (4.2%) had previous cardiac surgery, and 155 (13%) were on dialysis. 322 (27%) had bicuspid valves and 28 (2.3%) had infective endocarditis. 651 (54%) had aortic stenosis, 289 (24%) aortic regurgitation, and 87 (7.2%) mixed. 546 (46%) underwent concomitant procedures. Clinical outcomes, echocardiograms, and follow-up data were collected and analyzed for valve performance, and time-to-event analyses were performed for reoperation and mortality. 5023 patient-years of follow-up were available for analysis, with 50% of patients followed >3.2 years and 10% >9 years.

**Results:** Mean cardiopulmonary bypass and aortic clamp times for isolated Ozaki procedures were 151 ± 37 and 105 ± 29 minutes, respectively. Thirty-day mortality was 1.7% (n=20), new stroke 14 (2.6%), new dialysis 41(4.0%), and permanent pacemaker implantation 18 (1.5%). At 6 months, 5 years, and 10 years, peak/mean aortic valve gradients were 14.0/7.4, 15.5/8.0 and 15.5/8.2 mmHg, respectively, and ≥moderate regurgitation was 0.30%/2.9%/6.6%. Left ventricular mass index decreased from 141 ± 52 g/m^2^ preoperatively to 100 ± 1.1 g/m^2^ at 6 months and 90 ± 1.8 g/m^2^ at 10 years. At 10 years, freedom from reoperation was 91.2% and survival 75%.

**Conclusion:** The Ozaki procedure creates good aortic valves with stable low gradients. Regurgitation increased over time, but risk of reoperation was low, supporting continued use.

## INTRODUCTION

Aortic valve disease in younger patients poses unique challenges due to early structural deterioration and risk of reoperation with bioprosthetic valves, need for lifelong anticoagulation and its associated risk of bleeding and thromboembolic events with mechanical valves, and lack of durable repair options for stenotic valves. Although advent of valve-in-valve transcatheter aortic valve replacement (TAVR) broadens the lifetime management options available, long-term outcomes are unknown and subsequent open operations may prove increasingly challenging and risky.^1, 2^

In 2007, Ozaki and colleagues described and developed the tools for a new reproducible technique to create aortic valve cusps from glutaraldehyde-treated autologous pericardium, obviating the need for a prosthetic valve. Early results of the first 404 patients were reported in 2014, including stich-by-stich details of the operation.^3, 4^ Ozaki more recently reported mid-term results of 850 patients in 2017, demonstrating excellent outcomes and hemodynamics with a mean follow-up of 4 years.^5^ Since then, there has been a growing interest in this procedure, and other centers have published smaller series confirming reproducibility, safety, and early valve function.^6–9^ Long-term valve performance and durability remain to be proven, however, and at this stage, only Professor Ozaki’s pioneering series can provide initial insights.

In collaboration with Cleveland Clinic and Professor Ozaki’s colleagues, this study aimed to analyze the longer-term (up to 15 years) outcomes of the Ozaki procedure with focus on valve performance, need for reintervention, and survival by studying Ozaki’s series of patients.

## PATIENTS AND METHODS

### Patients

From April 2007 to May 2021, 1196 consecutive Ozaki procedures, 643 isolated and 553 with concomitant procedures, were performed at Toho University Ohashi Medical Center (Figure E1). During that period, fewer than 10 prosthetic valve implants were performed at Toho University. Use of the data was approved by the Toho University Institutional Review Board (#H22067, approved November 22, 2022), with patient consent waived. Data were provided for analysis by Professor Ozaki under a data use agreement with Cleveland Clinic.

### Ozaki Procedure

The procedure was performed with autologous pericardium in 95% of patients, but bovine pericardium (4%) and equine pericardium (1.2%) were used in a few cases, such as for reoperation or a minimally invasive approach. The excised autologous pericardium is stretched on a plate and tanned for 10 minutes with 0.6% glutaraldehyde solution, then rinsed in normal saline 3 times for 6 minutes. Size of the new cusps is determined by the intercommissural distances measured using Ozaki sizers (Tokyo Research Center for Advanced Surgical Technology Co., Ltd. Tokyo). Using the template, designed to create appropriate shape and generous coaptation, 3 cusps are drawn and cut from the treated and dried pericardium. Cusps are individually sutured to the anulus with 4-0 monofilament running suture. All patients were prescribed aspirin for 6 months after discharge. Technical modifications made by Professor Ozaki over the study period included the following (Figure E1):

- Switching the orientation of the pericardium. Initially, the rough side was facing the left ventricle, leading to a postoperative consumptive thrombocytopenia, prompting reversal with the smooth side facing the left ventricle (Case 19, January 2008).
- Adding a 5-mm “wing” extension to the pericardial cusps for more secure commissural fixation to prevent aortic regurgitation (AR) (Case 291, January 2011).
- Equal tricuspidization with implant of 3 equal-sized or 1-size-different cusps by adjusting the position of commissures as needed in patients with bicuspid and unicuspid valves. (Case 513, November 2012). This was done to minimize asymmetric cusp movement and potentially decrease risk of infective endocarditis.^5^
- New template and sizers were introduced to reduce excess coaptation height and facilitate future TAVR (Case 1147, July 2020).

### Endpoints

Endpoints included postoperative in-hospital morbidity and operative mortality, defined as for the Society of Thoracic Surgeons National database,^10^ postoperative follow-up valve hemodynamic metrics – peak and mean gradient – AR grade, left ventricular (LV) mass index assessed by transthoracic echocardiography, time-related aortic valve reintervention, and long-term mortality.

### Echocardiographic Follow-up

AR grade (none, mild, moderate, severe), mean gradient, and LV mass index were assessed on serial postoperative echocardiograms performed at approximately 1 week, 1 month, and every 6 months thereafter (Figure E2). A total of 5563 echocardiograms were available for 1050 patients (88% of study cohort). Median follow-up time was 2.8 years, with 556 (10% of echocardiographic studies) obtained after 7 years, and 80 patients had echocardiogram measurements available after 10 years (Figure E2). All longitudinal measurements were censored at aortic valve reoperation.

### Patient Follow-up for Postoperative Events

Systematic anniversary follow-up was used to assess reoperation events and vital status. For reoperation, 4968 patient-years of follow-up were available for analyses. Median follow-up was 3.2 years, with 25% followed more than 6.7 years, 10% more than 9 years, and 5% more than 10.5 years (Figure E3).

### Data Analysis

Statistical analyses were performed using SAS version 9.4 (SAS Institute, Cary, NC) and R version 3.6.0 (R Foundation for Statistical Computing, Vienna, Austria). Continuous variables are summarized as mean ± standard deviation or as equivalent 15th, 50th (median), and 85th percentiles when distribution of values was skewed. Categorical data are summarized by frequencies and percentages. Confidence intervals for longitudinal estimates used a bootstrap percentile method to obtain 68% confidence bands (equivalent to ±1 standard error) and the delta method for time-related events. A type I error of .05 was used to assess statistical significance.

#### Echocardiographic Analysis

To assess the temporal trend of individual grades of postoperative AR (ordinal longitudinal data), follow-up transthoracic echocardiograms were analyzed longitudinally for pattern of change across time using a nonlinear, multiphase, mixed-effects cumulative logit regression model.^11^ Prevalence of each AR grade over time was estimated by averaging patient-specific profiles. Note that because only 15 echocardiogram studies reported severe AR, it was combined with moderate AR grade. A multiphase, nonlinear, mixed-effects regression model was used similarly to estimate the temporal trend of postoperative mean gradient and left ventricular mass index (continuous longitudinal data).^12^ The models were implemented using PROC NLMIXED (SAS).

To identify variables related to AR grade, we used Boostmtree in R. This boosting approach is based on a marginal model for modeling longitudinal data.^13^ For this, we considered variables listed in Appendix E1. Missing data in the covariates were imputed “on the fly” as a part of growing the forest object.^14^ Variable importance was used to hierarchically order variables in relation to predicted AR,^15^ which can separate an overall effect of covariates into a main effects and a covariate–time interaction effect. One of main objectives of this machine learning approach is to visualize the relation of covariables to AR grade without model assumptions. For this, we used partial dependency plots, which describe the risk-adjusted relationship between the covariate of interest and AR grade by integrating out the effect of all other covariates.^16^

#### Time-to-Event Analysis

Survival and freedom from reoperation were estimated nonparametrically by the Kaplan-Meier method and parametrically using a multiphase hazard method.^17^

We performed random forests for survival (time-related events) to identify factors associated with time-related reoperation. Random forests is a nonparametric, statistical ensemble method that uses all variables. It makes no distributional or functional (linear or nonlinear) or interaction-effects assumptions about covariate relationships to the response.^19^ Random forest analyses were carried out using the randomForestSRC package in R.^18^ For these analyses, a forest was grown using 5000 regression trees with the variables listed in Appendix E1. Missing data, estimation of variable importance, and partial dependence plots are handled as described above.

To identify risk factors for death, multivariable analysis was performed in the multiphase hazard function^17^ domain using the variables listed in Table E1. Prior to multivariable analysis, 5-fold multiple imputation^20^ using a multivariate imputation by chained equations (MICE) method was used to impute missing data (SAS PROC MI). Variable selection, with a *P*-value criterion for retention of variables in the model of .05, used bagging (bootstrap aggregation).^19^ All variables with bootstrap reliability of 50% or greater were retained as risk factors in the analysis. Then following Rubin,^20^ we combined estimates from the 5 models using these same identified risk factors (SAS PROC MIANALYZE) to yield final regression coefficient estimates, the variance–covariance matrix, and *P* values. As a complementary analysis, we also performed random forests for survival to identify factors associated with risk of death as described above.

To estimate intrinsic risk of valve reoperation in the absence of mortality, a competing-risks analysis was performed by considering the mutually exclusive outcomes of reoperation and death before reoperation. A common interval was defined for analysis as the earliest of either death or reoperation, or censored event for patients still alive but having no reoperation. Freedom from each event was the estimated by the nonparametric product limit method.^21^ Variances of the estimates were based on the Greenwood formula.^21^ The instantaneous risk (hazard function) for each competing event was estimated by a parametric method.^17^ Consequences of the independent, simultaneous operative transition rates (hazard functions) from the followed living patients without reoperation into either death or reoperation were calculated by integrating the parametric equations.

## RESULTS

All patients were Japanese with a mean age of 68 years (range 11 to over 90 years); 712 (60%) were male, 50 (4.2%) had previous cardiac surgery, and 155 (13%) were on dialysis (Table 1). Average body mass index was 22 kg/m^2^, and 201 (17%) had diabetes. Bicuspid aortic valves were present in 322 (27%), and 28 (2.3%) had infective endocarditis. Pure aortic stenosis (AS) was present in 651 (54%), 289 (24%) had pure AR, and 87 (7.2%) had mixed AS and AR. Concomitant procedures were performed in 551 (46%) (Table E1).

**Table 1:**
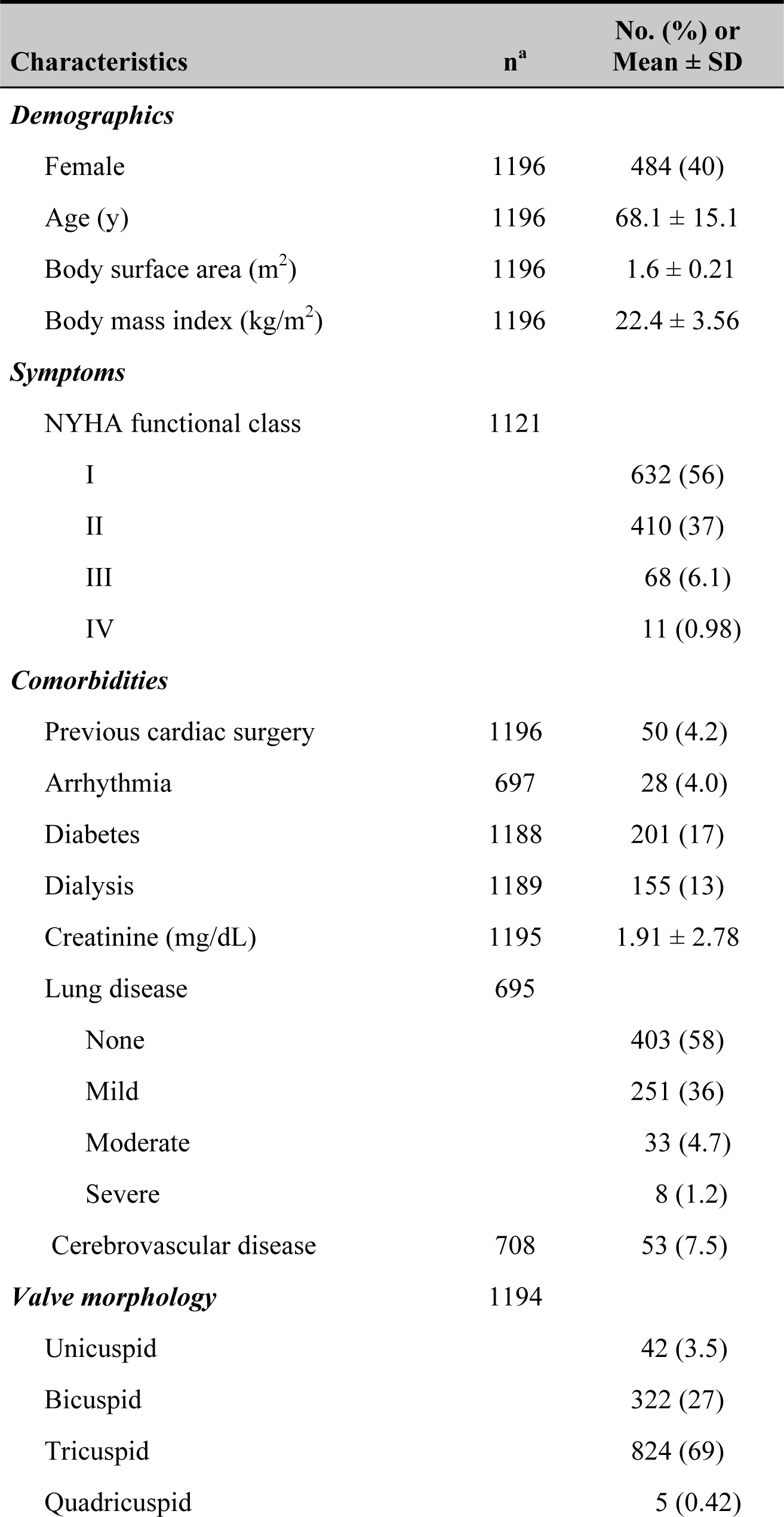
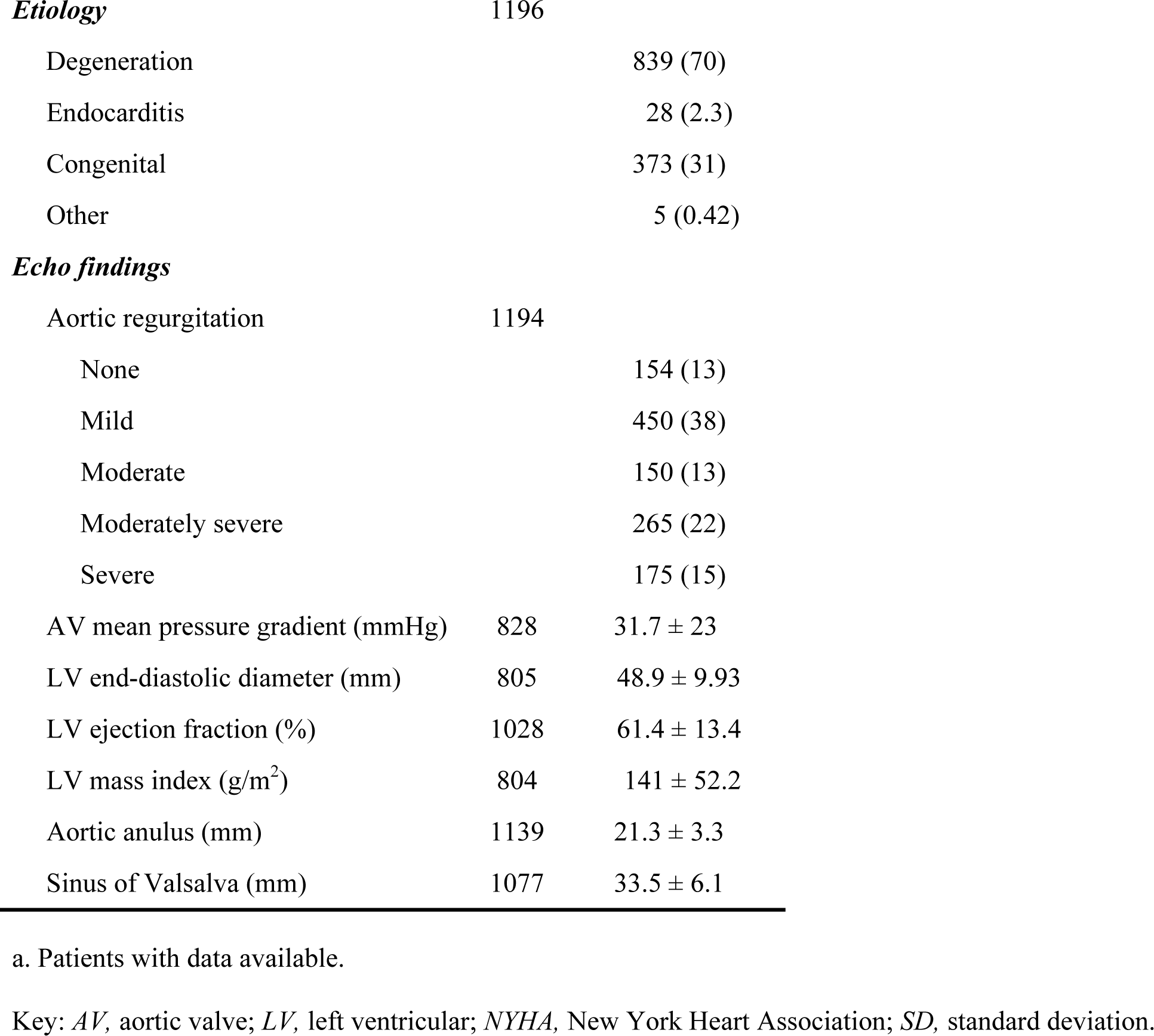
Preoperative Patient Characteristics (n=1196)

| Characteristics | n<sup>a</sup> | No. (%) or Mean ± SD |
| --- | --- | --- |
| ***Demographics*** |  |  |
| Female | 1196 | 484 (40) |
| Age (y) | 1196 | 68.1 ± 15.1 |
| Body surface area (m<sup>2</sup>) | 1196 | 1.6 ± 0.21 |
| Body mass index (kg/m<sup>2</sup>) | 1196 | 22.4 ± 3.56 |
| ***Symptoms*** |  |  |
| NYHA functional class | 1121 |  |
| I |  | 632 (56) |
| II |  | 410 (37) |
| III |  | 68 (6.1) |
| IV |  | 11 (0.98) |
| ***Comorbidities*** |  |  |
| Previous cardiac surgery | 1196 | 50 (4.2) |
| Arrhythmia | 697 | 28 (4.0) |
| Diabetes | 1188 | 201 (17) |
| Dialysis | 1189 | 155 (13) |
| Creatinine (mg/dL) | 1195 | 1.91 ± 2.78 |
| Lung disease | 695 |  |
| None |  | 403 (58) |
| Mild |  | 251 (36) |
| Moderate |  | 33 (4.7) |
| Severe |  | 8 (1.2) |
| Cerebrovascular disease | 708 | 53 (7.5) |
| ***Valve morphology*** | 1194 |  |
| Unicuspid |  | 42 (3.5) |
| Bicuspid |  | 322 (27) |
| Tricuspid |  | 824 (69) |
| Quadricuspid |  | 5 (0.42) |
| ***Etiology*** | 1196 |  |
| Degeneration |  | 839 (70) |
| Endocarditis |  | 28 (2.3) |
| Congenital |  | 373 (31) |
| Other |  | 5 (0.42) |
| ***Echo findings*** |  |  |
| Aortic regurgitation | 1194 |  |
| None |  | 154 (13) |
| Mild |  | 450 (38) |
| Moderate |  | 150 (13) |
| Moderately severe |  | 265 (22) |
| Severe |  | 175 (15) |
| AV mean pressure gradient (mmHg) | 828 | $31.7 \pm 23$ |
| LV end-diastolic diameter (mm) | 805 | $48.9 \pm 9.93$ |
| LV ejection fraction (%) | 1028 | $61.4 \pm 13.4$ |
| LV mass index ($\text{g}/\text{m}^2$) | 804 | $141 \pm 52.2$ |
| Aortic anulus (mm) | 1139 | $21.3 \pm 3.3$ |
| Sinus of Valsalva (mm) | 1077 | $33.5 \pm 6.1$ |

a. Patients with data available.
Key: AV, aortic valve; LV, left ventricular; NYHA, New York Heart Association; SD, standard deviation.

### Valve Hemodynamic Performance

The percentage of patients with moderate or severe postoperative AR gradually increased over time, from 0.59% at 1 year to 6.6% at 10 years (Figure 1). Patients who had equal tricuspidization had a lower risk of developing AR after 5 years, and patients younger than 70 had a slightly higher risk of developing long-term AR. Bicuspid valve and large sinus diameter were not associated with increased risk of AR (Figure E4). Five- and 10-year risk of moderate or greater AR decreased during the earlier part of the series but increased slightly after 2012 (Figure 2).

**Figure 1:**
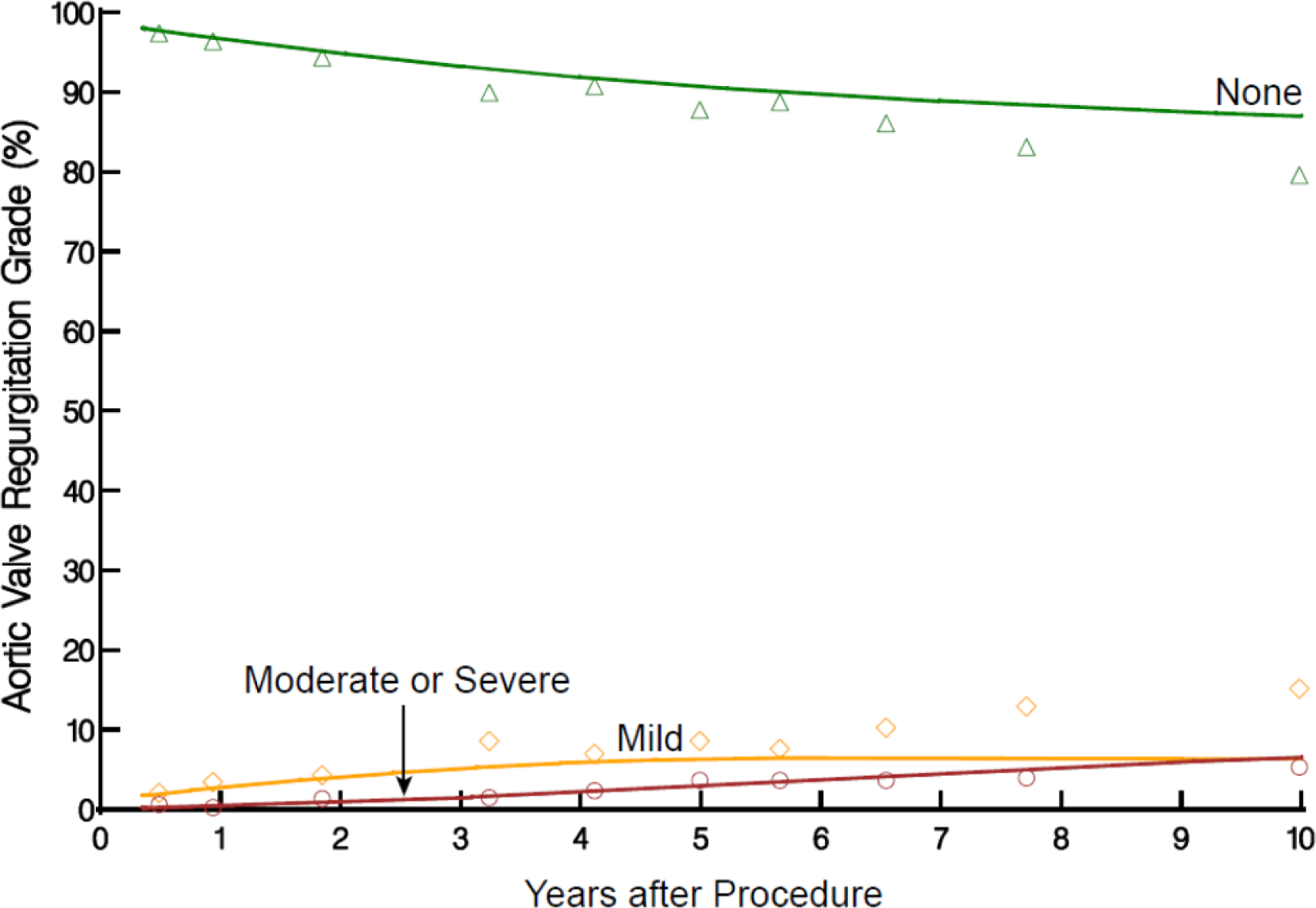
Temporal trend of postoperative aortic regurgitation (AR) grade after Ozaki procedure. Solid lines represent parametric estimates of percentage of patients in each postoperative AR grade after surgery. Symbols represent data grouped (without regard to repeated measurements) within the time frame to provide a crude verification of model fit.

**Figure 2:**
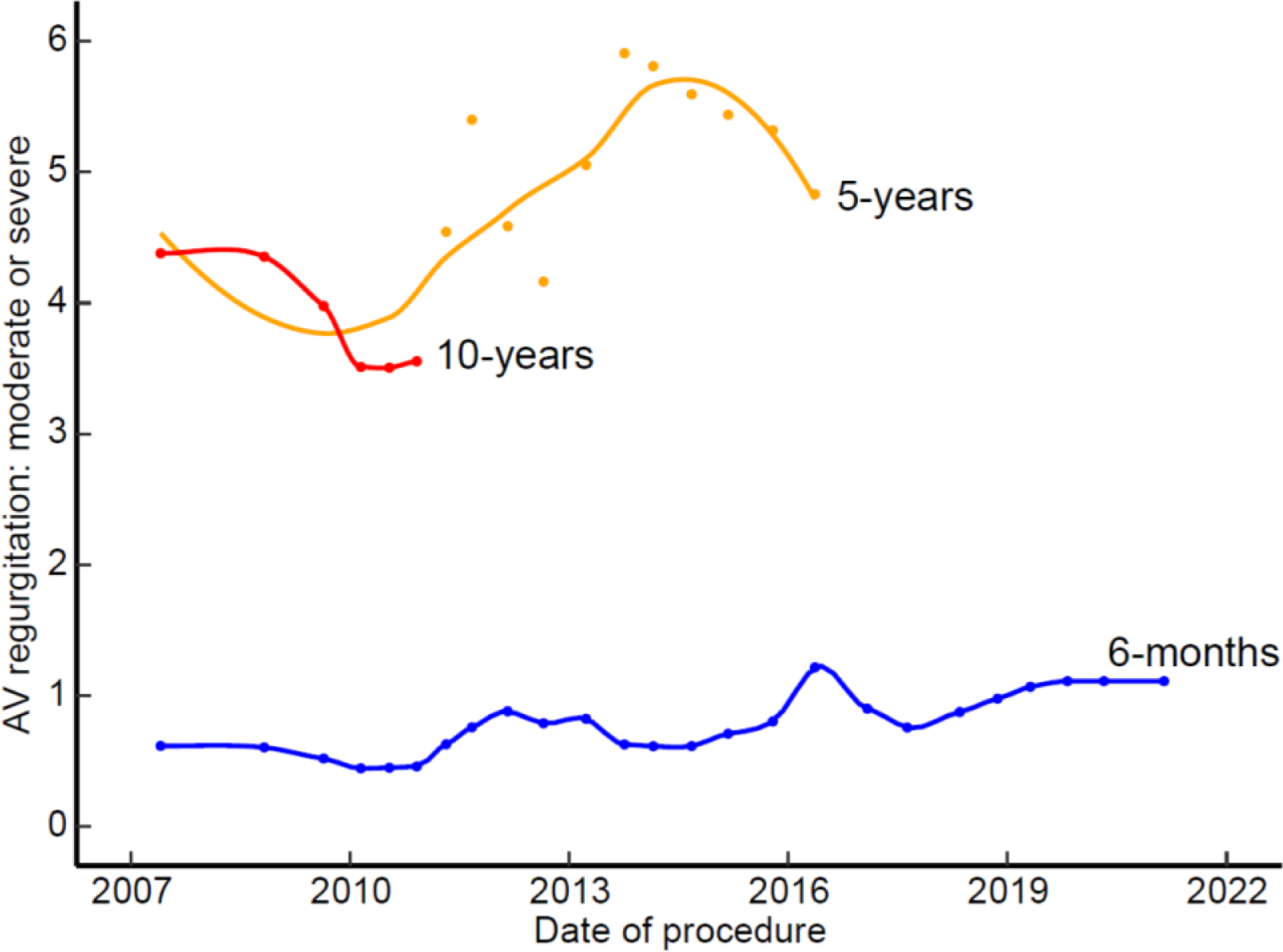
Association between date of surgery and likelihood of moderate or severe aortic regurgitation (AR) after surgery. Partial dependency plot of predicted prevalence of moderate or severe AR and date of surgery. Symbols are risk-adjusted predicted 6-month (blue line), 5-year (orange line), and 10-year (red line) probability of moderate or severe AR at different values of selected continuous covariables. Solid lines are smoothed loess lines of the symbols.

Mean aortic valve gradient was 7.4 mmHg at 6 months, 8.1 mmHg at 3 years, and 8.2 mmHg at 10 years (Figure 3). Use of non-autologous pericardium to create the cusps, smaller cusp size, smaller anulus size, and higher creatinine were associated with higher postoperative mean gradients, but younger age was not (Figure E5).

**Figure 3:**
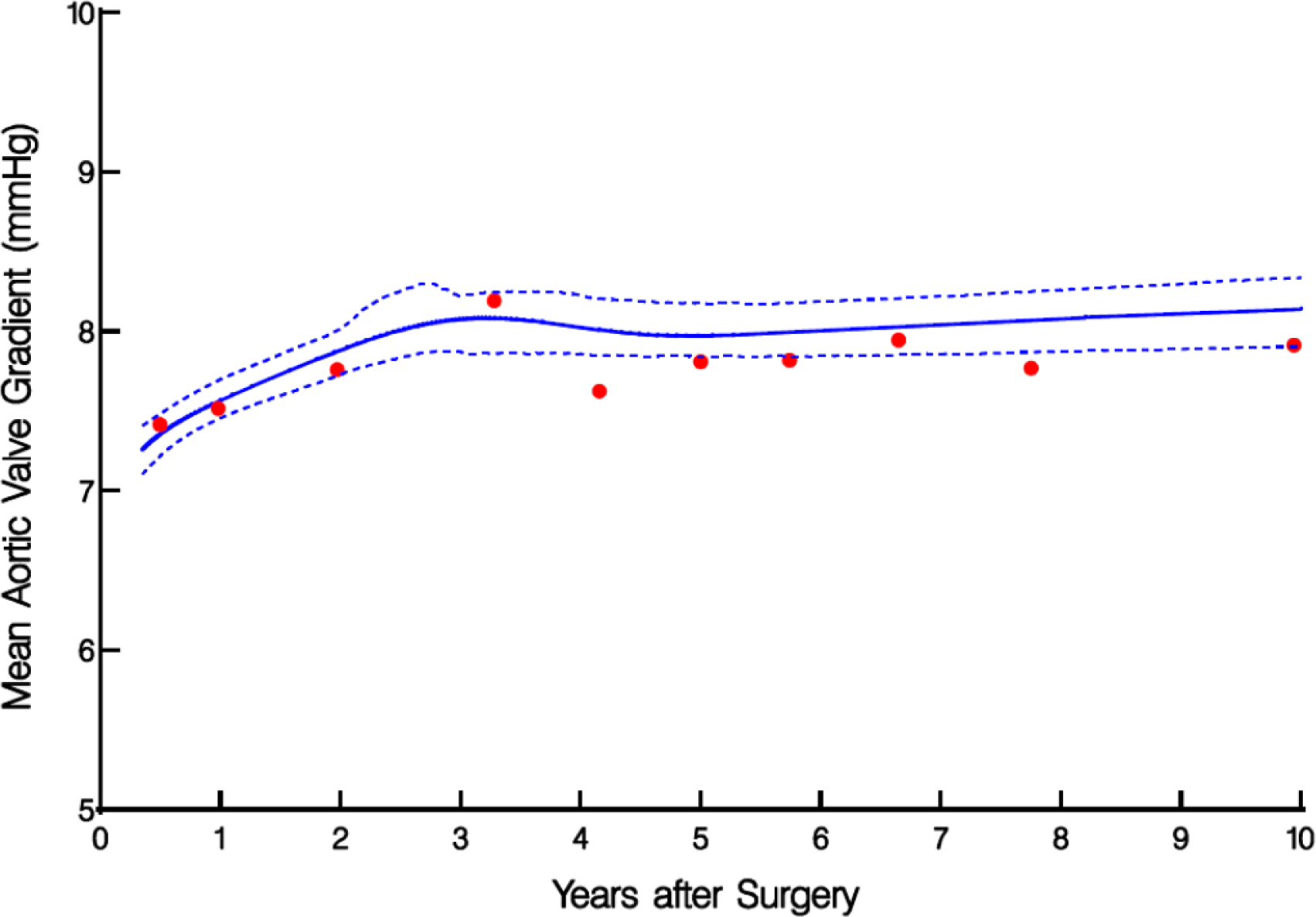
Temporal trend of aortic valve (AV) mean gradient after Ozaki procedure. Solid lines represent parametric estimates of AV mean gradient after the surgery, enclosed within 68% confidence intervals. Symbols represent data grouped (without regard to repeated measurements) within the time frame to provide a crude verification of model fit.

LV mass index gradually decreased over time, from 141 g/m^2^ preoperatively, to 100 g/m^2^ at 6 months and about 90 g/m^2^ by 10 years (Figure 4).

**Figure 4:**
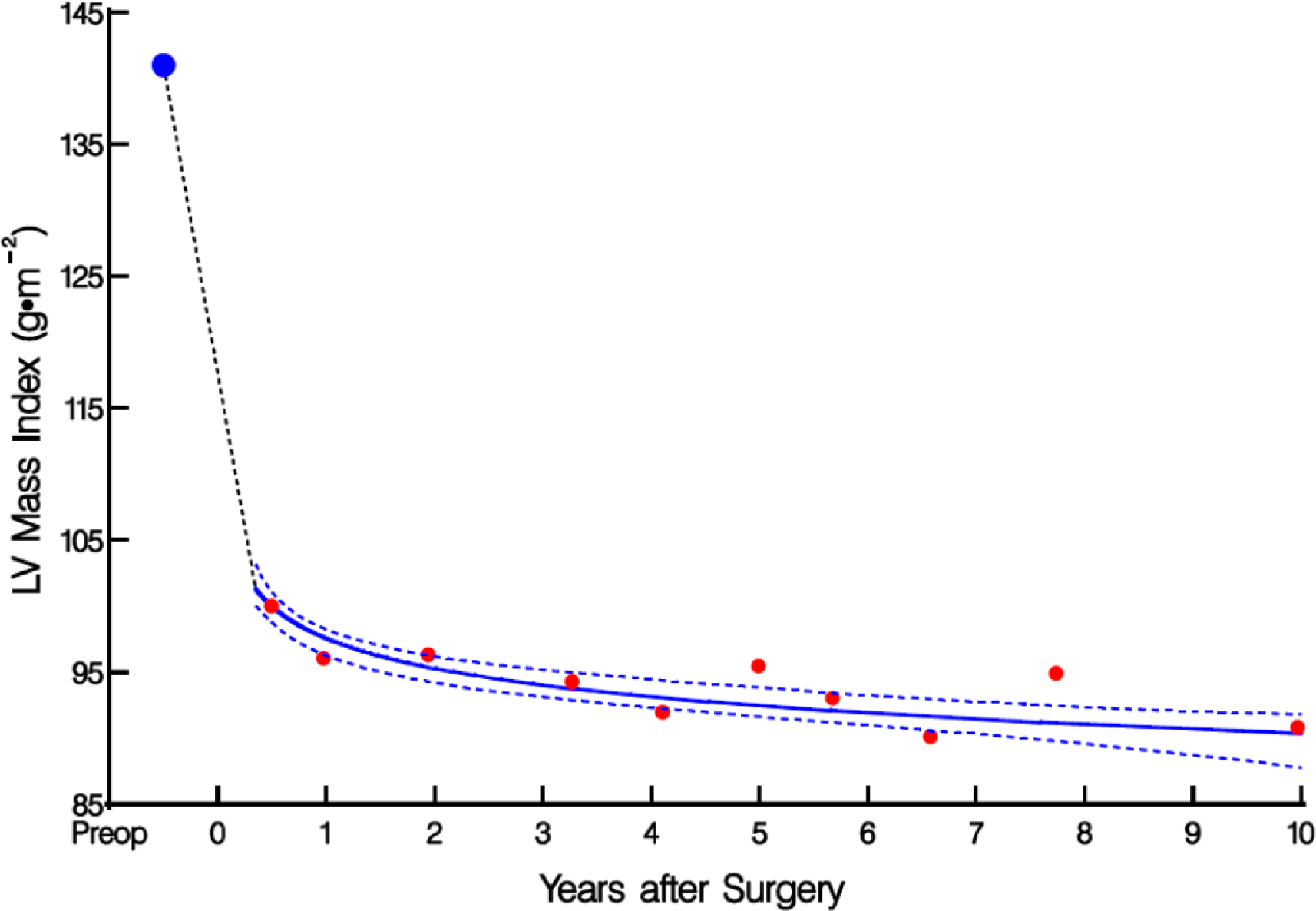
Temporal trend of LV mass index after Ozaki procedure. Solid lines represent parametric estimates of LV mass index after the surgery. Format is as in Figure 3.

### Aortic Valve Reoperation

During follow-up, there were 38 reoperations, 17 for endocarditis, 18 for degeneration (AR in 15, AS in 3), 2 for suture tear, and 1 for anular dilatation. There were no reoperations in patients with non-autologous Ozaki cusps. Seven reoperations were for endocarditis after equal tricuspidization. Actuarial occurrence (Kaplan-Meier) of aortic valve re-replacement was 0.4% at 1 year, 3.4% at 5 years, and 8.8% at 10 years (Figure 5). Cusp size difference >6, sinus diameter >40 mm, young age, bicuspid valve, and dialysis were factors associated with higher risk of reoperation (Figure E6). The cumulative incidence of patients experiencing reoperation was estimated to be 0.35%, 3.1%, and 7.4% at 1, 5, and 10 years, respectively. Conditional probability of reoperation was slightly higher than the actuarial estimates at 0.37%, 3.6%, and 9.6% at the same time points (Figures 6 and E7).

**Figure 5:**
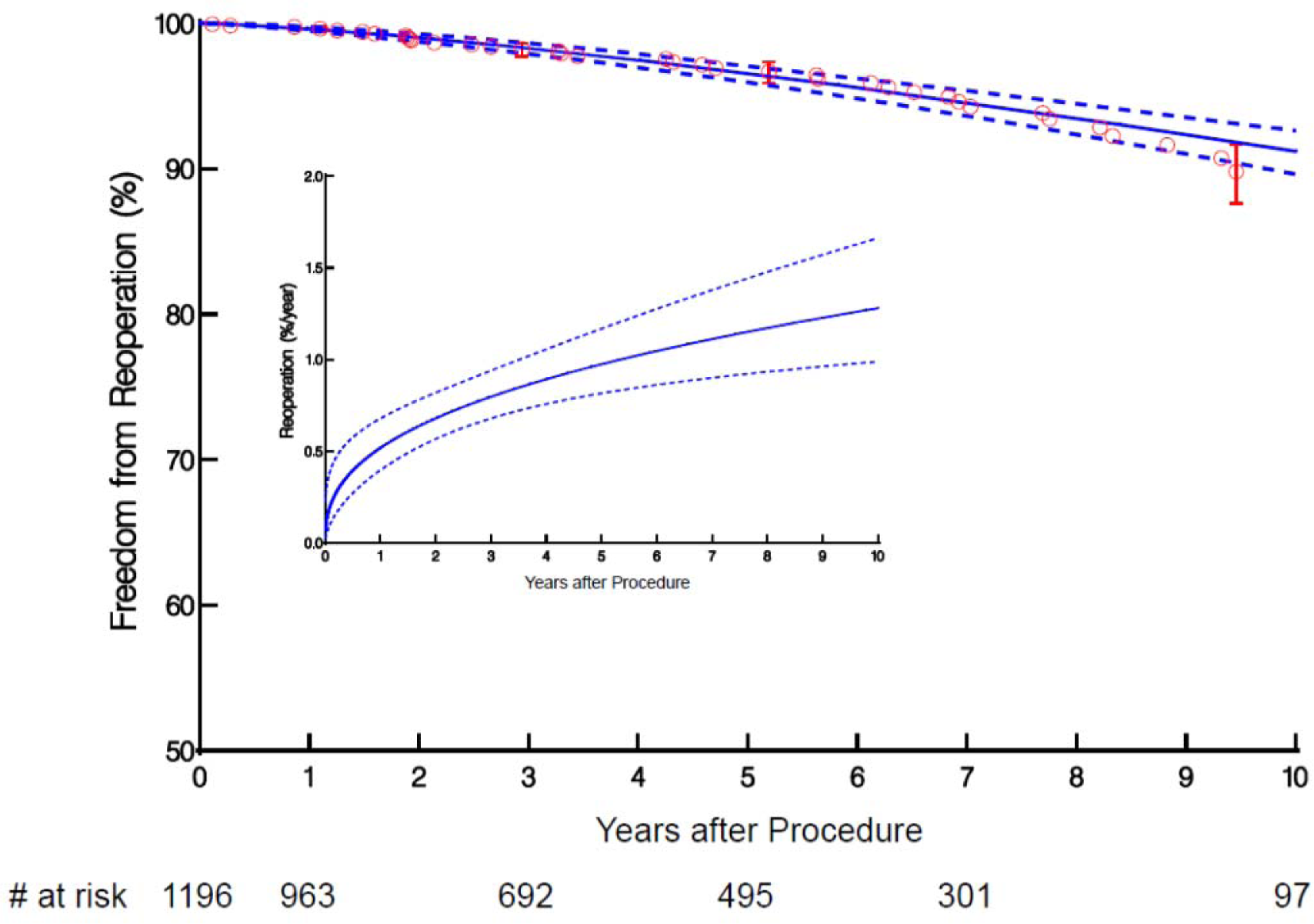
Freedom from reoperation after Ozaki procedure. Solid line is parametric estimate enclosed within a 68% confidence interval. Kaplan-Meier nonparametric estimates are given by the respective symbols enclosed within the 68% confidence bars equivelent to ±1 standard error at selected time points. Number of patients at risk at different time points are provided under the horizontal axis. **Inset**: Instantaneous risk. Solid lines are parametric estimates enclosed within a 68% confidence interval.

**Figure 6:**
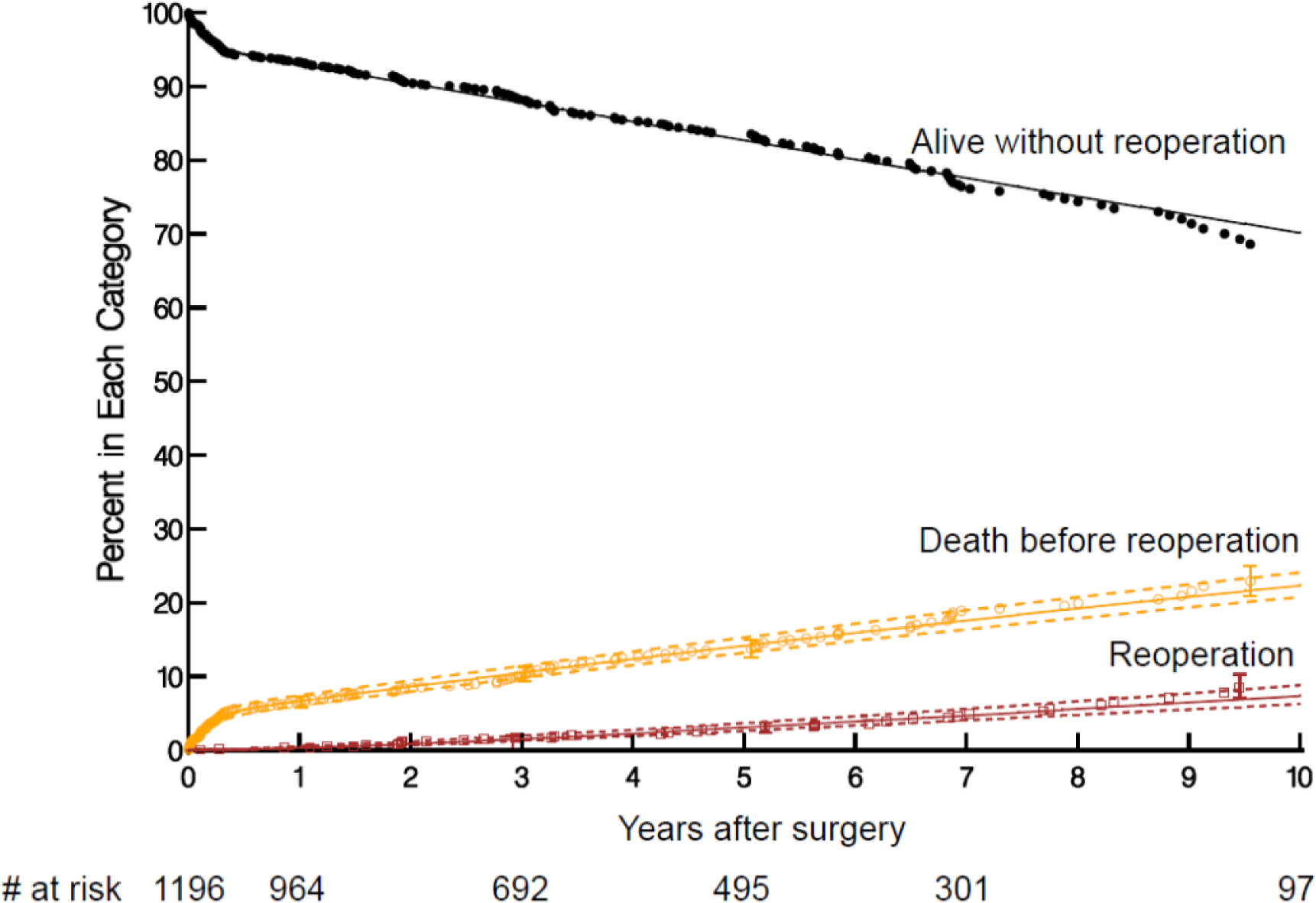
Probability of occurrence of competing events after Ozaki procedure (cumulative incidence function). Solid lines represent parametric estimates enclosed within 68% confidence bands equivalent to ±1 standard error. Symbols are nonparametric estimates with 68% confidence bars. At each time point, “Alive without reoperation” (some would call this “event-free survival”), “Death before reoperation,” and “reoperation” add to 100%.

### In-Hospital Morbidity/Mortality

Thirty-one patients (2.6%) had a stroke, 41 (4%) developed renal failure requiring dialysis, and 18 (1.5%) developed complete heart block requiring a pacemaker (Table E2). Three of 643 (0.5%) patients undergoing an isolated Ozaki procedure required a pacemaker, compared with 15 of 553 (2.7%) undergoing concomitant procedures. Thirty-day mortality was 1.7%.

### Late Mortality

There were 166 deaths during follow-up, 19 cardiac, 124 noncardiac, and 23 unknown. An early high-risk phase was followed by a slightly increasing late risk of death of 2% to 3% per year after 1 year (Figure 7). Survival at 6 months, and 1, 3, 5, and 10 years after the Ozaki procedure was 94.4%, 93.4%, 89.8%, 85.7%, and 75.2%, respectively, which is comparable to the age-sex– matched Japanese population norm. Older age, kidney failure, and cusp size <21 were associated with increased risk of death (Table E3 and Figures E8 and E9).

**Figure 7:**
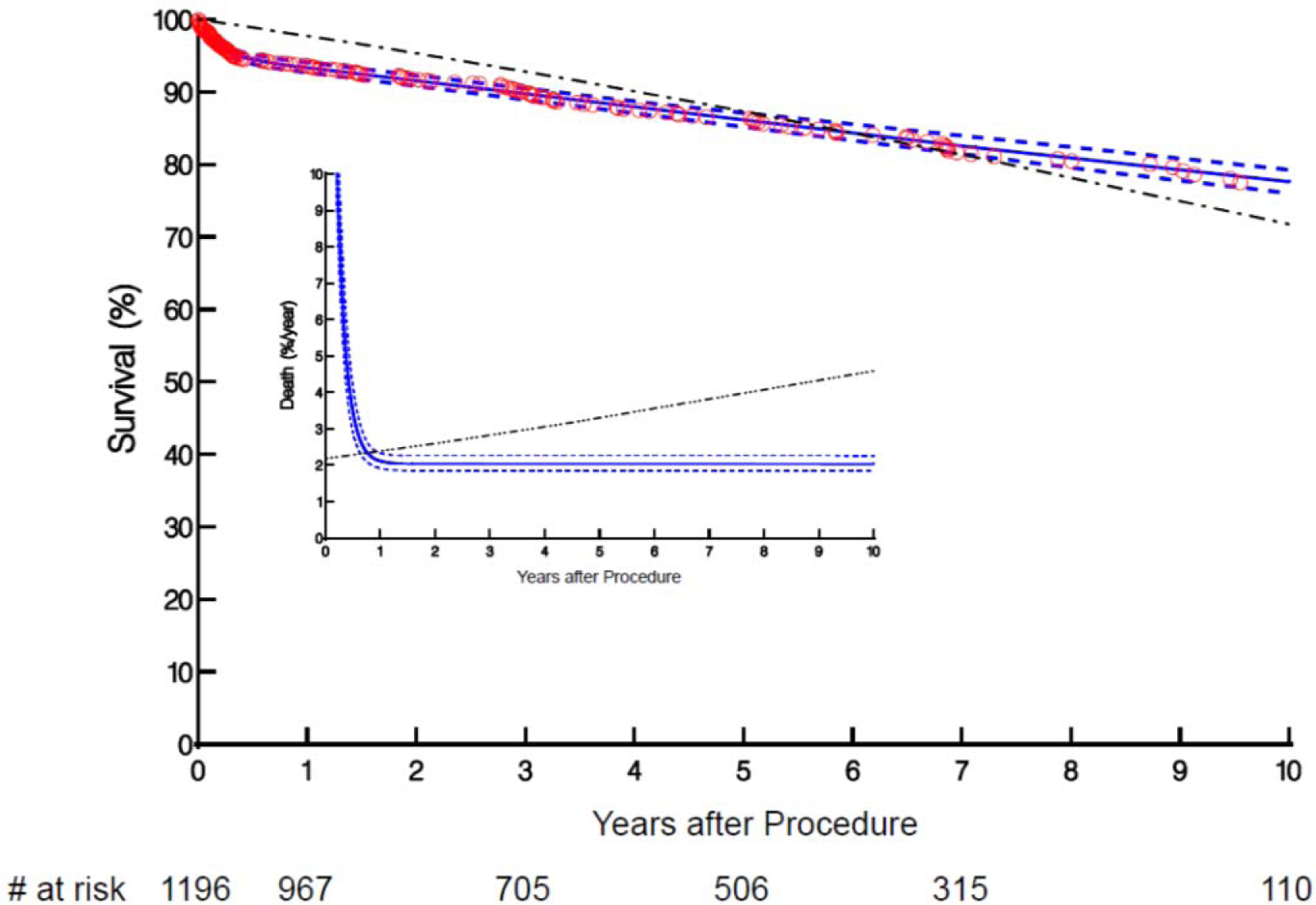
Survival after Ozaki procedure. Solid line is parametric estimate enclosed within a 68% confidence interval. The Kaplan-Meier nonparametric estimates are given by the respective symbols enclosed within the 68% confidence bars equivelent to ±1 standard error at selected time points. Dash-dot-dot line depicts the corresponding expected survival of an age-sex– matched Japanese population. Number of patients at risk at different time points are provided below the horizontal axis. **Inset**: Instantaneous risk of death. Solid lines are parametric estimates of instantaneous risk of death enclosed within a 68% CI. Dash-dot-dot line depicts the age-sex– matched Japanese population’s expected life.

## DISCUSSION

### Principal Findings

Our study adds the following to the existing literature:

- Longitudinal valve performance was excellent up to 10 years
- Ozaki valves had

`o` Low gradients
`o` Low but slightly increasing risk of AR over time
- Reverse LV remodeling continued up to 10 years
- Risk of reoperation was low
- Survival was excellent and comparable to that of the general Japanese population

### Advantages of the Ozaki Procedure

1. Sizer, template, instructions for preparing the pericardium, and detailed stitch-by-stitch instructions make the operation very reproducible.
2. The Ozaki valve preserves anulus mobility and has a large effective orifice area (EOA) …
3. resulting in laminar flow and valve kinetics similar those of a native valve.
4. The large coaptation height minimizes cusp stress.
5. It is a single-valve operation that preserves the native aortic root, …
6. has no need for anticoagulation, …
7. has low risk of pacemaker implant, … and
8. avoids the cost of a replacement valve.
9. The use of autologous tissue is attractive to patients and surgeons.

### Valve Hemodynamics – Large Effective Orifice Area

Early results of the Ozaki procedure presented by multiple centers have shown excellent hemodynamic performance. Rosseykin and colleagues reported that patients who underwent the Ozaki procedure had lower gradients and larger valve area compared with those who received Hancock II and PERIMOUNT bioprostheses.^22^ Our recently published study similarly showed a lower gradient but higher long-term AR compared with the PERIMOUNT bioprosthesis.^23^ Standard bioprostheses or mechanical valves are rigid, which restricts the natural motion of the aortic anulus; the normal anulus expands in systole, which increases the EOA by 10% to 15%.^24^ Standard stentless valves have superior hemodynamics compared with stented prostheses, but the extra tissue required to support the valve limits the EOA, and durability is similar or even less than for stented bioprosthetic valves.^25–27^ A meta-analysis by Mylonas and colleagues, including 22 studies of 1891 patients who underwent Ozaki procedures, reported a postoperative mean gradient of 7.7 mmHg, and a mean EOA index of 2.1±0.5 cm^2^/m^2^ at discharge,^28^ which is larger than the Magna Ease (1.2 ± 0.25 cm^2^/m^2^)^29^ or Freestyle stentless valve (0.9–1.0 cm^2^/m^2^). ^30^ Iida and colleagues demonstrated effectiveness of the Ozaki procedure in patients with a small aortic anulus (average 20 ± 2.5 mm), with an EOA index of 1.2 ± 0.4 cm/m^2^. ^31^ In addition to the large EOA at rest, EOA increased with increased flow. Saisho and colleagues demonstrated in an ex-vivo study that EOA of the Ozaki valve was similar to that of the native aortic valve and substantially larger than that of bioprosthetic or mechanical valves. Whereas Ozaki valves had an increase in EOA similar to the native valve, prosthetic valves showed a weaker increase in EOA with increased flow.^32^ The flow pattern on 4-Dimensional CT showed laminar flow similar to that of native valves.^33^

### Aortic Regurgitation

There is a risk of AR with the Ozaki procedure. We previously demonstrated that this risk had a learning curve; 5-year risk of severe AR rapidly decreased over the first 300 cases.^23^ In an effort to address this issue, wing extension and equal tricuspidization were introduced: wing extension (November 2011) was added to improve commissure strength and equal tricuspidization (February 2012) to minimize asymmetric cusp movement by creating symmetrical neo-commissures in patients with non-tricuspid aortic valves (of note, equal tricuspidization also aimed to reduce the risk of infective endocarditis). Interestingly, our analysis revealed that the 5-year risk of AR decreased initially, then increased around the time these modifications were introduced. It is therefore unclear what effect these modifications have had on the risk of AR. During this period, surgeons in training were involved in portions of the procedures, but this increase may simply represent another learning phase of the new techniques. The 5-year risk of AR started to decrease again around 2016. Infective endocarditis was another important cause of AR, discussed below.

### Left Ventricular Reverse Remodeling

The Ozaki procedure had an immediate LV mass reduction of 30% at 6 months, followed by a continued reduction to 36% at 10 years, which is better than reports for stented bioprostheses or TAVR.^26, 34–38^ Despite the increase in long-term AR, improved EOA and aortic anulus expansion with increased cardiac output are likely explanations for the observed impressive reverse remodeling.

### Reoperation for Structural Valve Deterioration and Endocarditis

Freedom from reoperation was 91.2% at 10 years, comparable to the risk of reoperation after a Ross procedure: The German Ross registry reports freedom from reoperation at 12 years for both autografts and allografts of 88% with the subcoronary technique, 77% with root replacement, and 87% with root replacement with reinforcement.^39–41^ Ozaki valve kinetics mimic those of the native valve, as both valves open before the onset of blood flow through the valve, a result of expansion of the aortic root.^42^ This mechanism is thought to reduce cusp stress and degeneration.

Use of autologous tissue was presumed to prevent endocarditis, but half the reoperations were due to endocarditis, similar to other series of prosthetic valves.^43^ Previous reports of aortic valve replacement using autologous pericardium have reported similar increased risk of endocarditis.^44, 45^ In 2012, Ozaki initiated equal tricuspidization by creating neo-commissures to prevent uneven opening of the cusps, which he hypothesized might increase risk of endocarditis. However, there has been no significant difference in the risk of reoperation due to endocarditis before and after implementation of equal tricuspidization.

Although kidney failure was a risk factor for reoperation, few patients had calcific degeneration requiring reintervention among dialysis patients.^46^

### Use of Alternative Material

A few patients lacked sufficient useful autologous pericardium or preferred a minimally invasive approach and had their Ozaki procedures performed with bovine or equine pericardium. Although use of autologous tissue is one of the major attractions of the procedure, use of xenopericardium has several advantages: it enables a minimally invasive approach, can be used in reoperations, and saves operative time spent harvesting, preparing, tanning, and rinsing the pericardium. At this point, the small number of patients and short follow-up do not allow making recommendations, but so far there have been no reoperations. We noticed slightly higher gradients, however.

### Limitations

The main limitation of this study is that the majority of the Ozaki procedures were done by a single surgeon. Although the sizers, template, and detailed description make this operation reproducible, the generalizability of the results needs to be confirmed by multiple centers.

## CONCLUSIONS

The Ozaki procedure creates a good aortic valve with low stable gradients, acceptable risk of AR up to at least 10 years, excellent LV remodeling, and a low risk of reoperation. Although longer term follow up is needed, this study confirms the safety and effectiveness of the Ozaki procedure required to justify expanded use of the procedure. Further studies are needed for optimal patient selection and to compare hemodynamics and durability with commercially available prostheses and the Ross procedure.

## Data Availability

The method of the analysis will be made available from the corresponding author on request.

## NON-STANDARD ABBREVIATIONS AND ACRONYMS

AR: aortic regurgitation
LV: left ventricle
EOA: effective orifice area
TAVR: transcatheter aortic valve replacement

## Acknowledgments

The authors thank Tess Parry for editorial contributions and Brian Kohlbacher for audiovisual contributions.

## Supplemental [Online-Only] Material

**Table E1:**
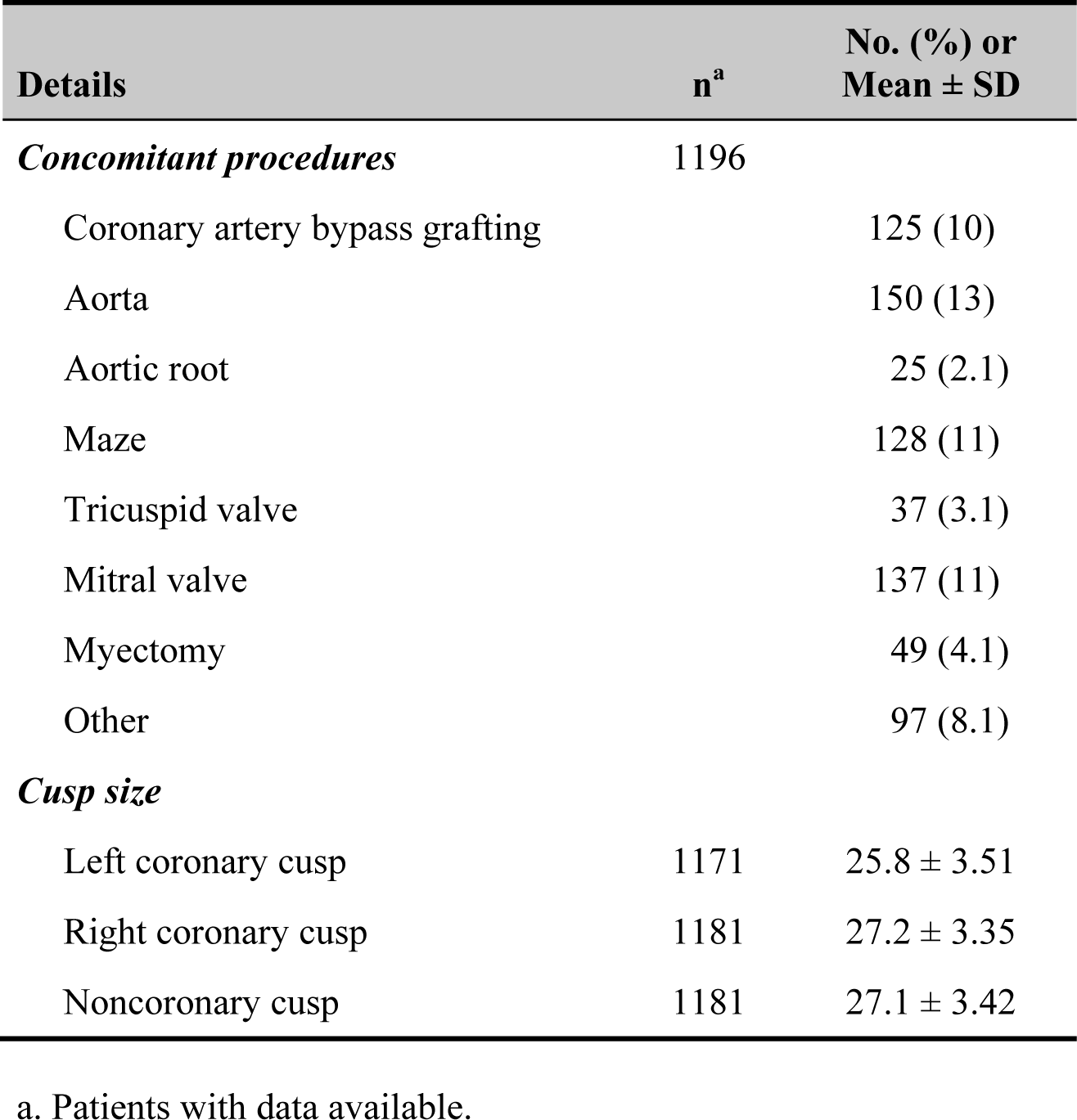
Operative Details (n=1196)

| Details | n<sup>a</sup> | No. (%) or Mean ± SD |
| --- | --- | --- |
| *Concomitant procedures* | 1196 |  |
| Coronary artery bypass grafting |  | 125 (10) |
| Aorta |  | 150 (13) |
| Aortic root |  | 25 (2.1) |
| Maze |  | 128 (11) |
| Tricuspid valve |  | 37 (3.1) |
| Mitral valve |  | 137 (11) |
| Myectomy |  | 49 (4.1) |
| Other |  | 97 (8.1) |
| *Cusp size* |  |  |
| Left coronary cusp | 1171 | 25.8 ± 3.51 |
| Right coronary cusp | 1181 | 27.2 ± 3.35 |
| Noncoronary cusp | 1181 | 27.1 ± 3.42 |

a. Patients with data available.

**Table E2:**
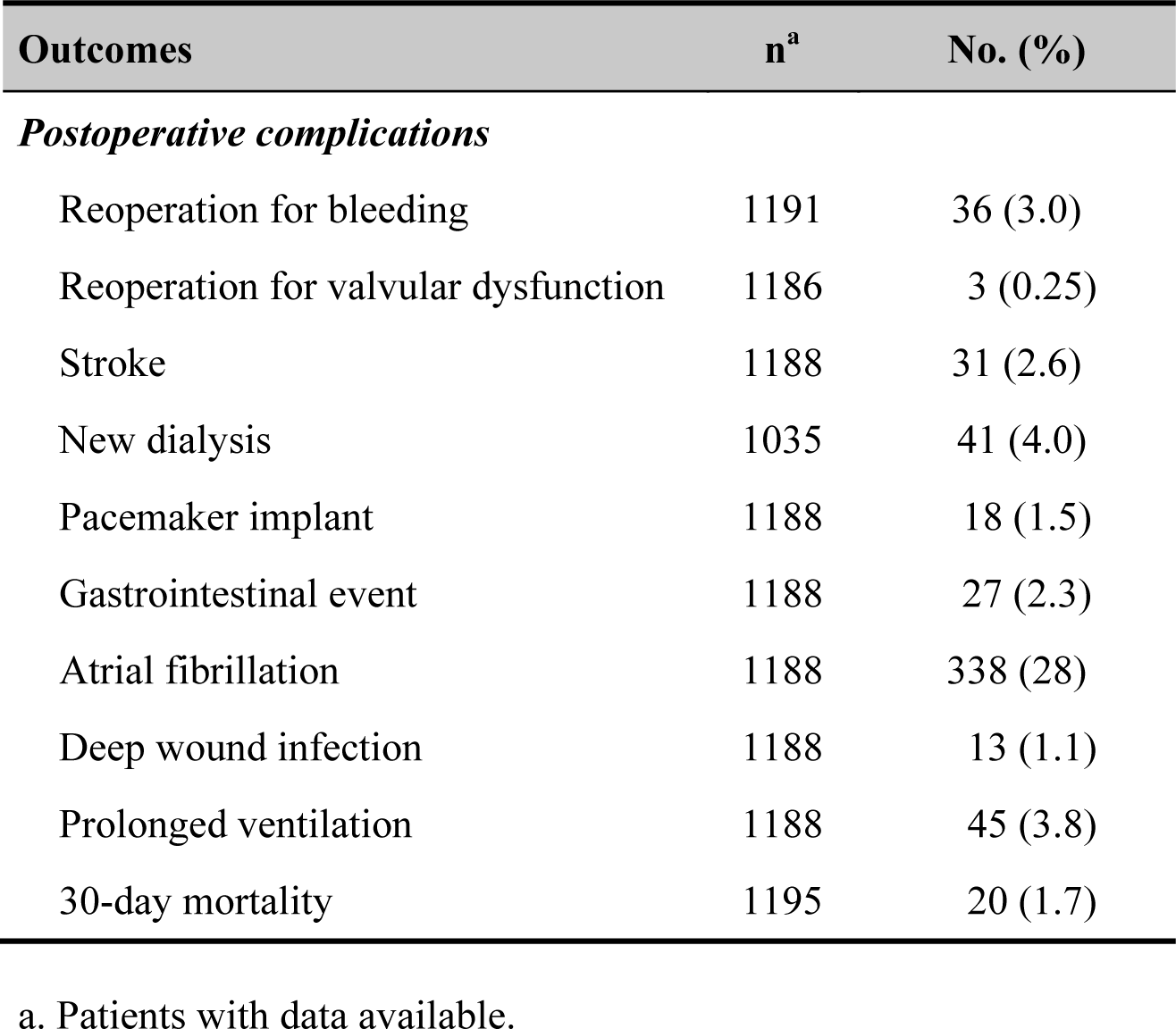
In-Hospital Outcomes (n=1196)

**Table E3:** Incremental Risk Factors for Death after Ozaki Procedure

| **Factor** | **Coefficient ± SE** | **P-Value** | **Reliability (%)<sup>a</sup>** |
| --- | --- | --- | --- |
| ***Early hazard phase*** |  |  |  |
| Older age<sup>b</sup> | 1.2 ± 0.24 | <.0001 | 87 |
| Higher creatinine<sup>c</sup> | -2.5 ± 0.40 | <.0001 | 90 |
| Concomitant MV procedure | 0.74 ± 0.34 | .03 | 85 |
| Tricuspidization (less risk) | -1.2 ± 0.35 | .0008 | 74 |
| ***Late hazard phase*** |  |  |  |
| Older age<sup>d</sup> | 0.81 ± 0.13 | <.0001 | 99 |
| Higher creatinine<sup>e</sup> | 0.63 ± 0.095 | <.0001 | 97 |
| Lower LV ejection fraction<sup>f</sup> | 0.42 ± 0.21 | .044 | 63 |
| Any concomitant procedure | 0.56 ± 0.20 | .005 | 91 |

a. Bagging reliability: Proportion of 1000 bootstrap analyses in which this variable was retained with $P < .05$ .
b. Exp[age/50], exponential transformation.
c. [1/creatinine], inverse transformation.
d. Exp[age/50], exponential transformation.
e. Log[creatinine], logarithmic transformation.
f. [50/LV ejection fraction], inverse transformation.
Key: *MV*, mitral valve; *LV*, left ventricular; *SE*, standard error.

**Figure E1:**
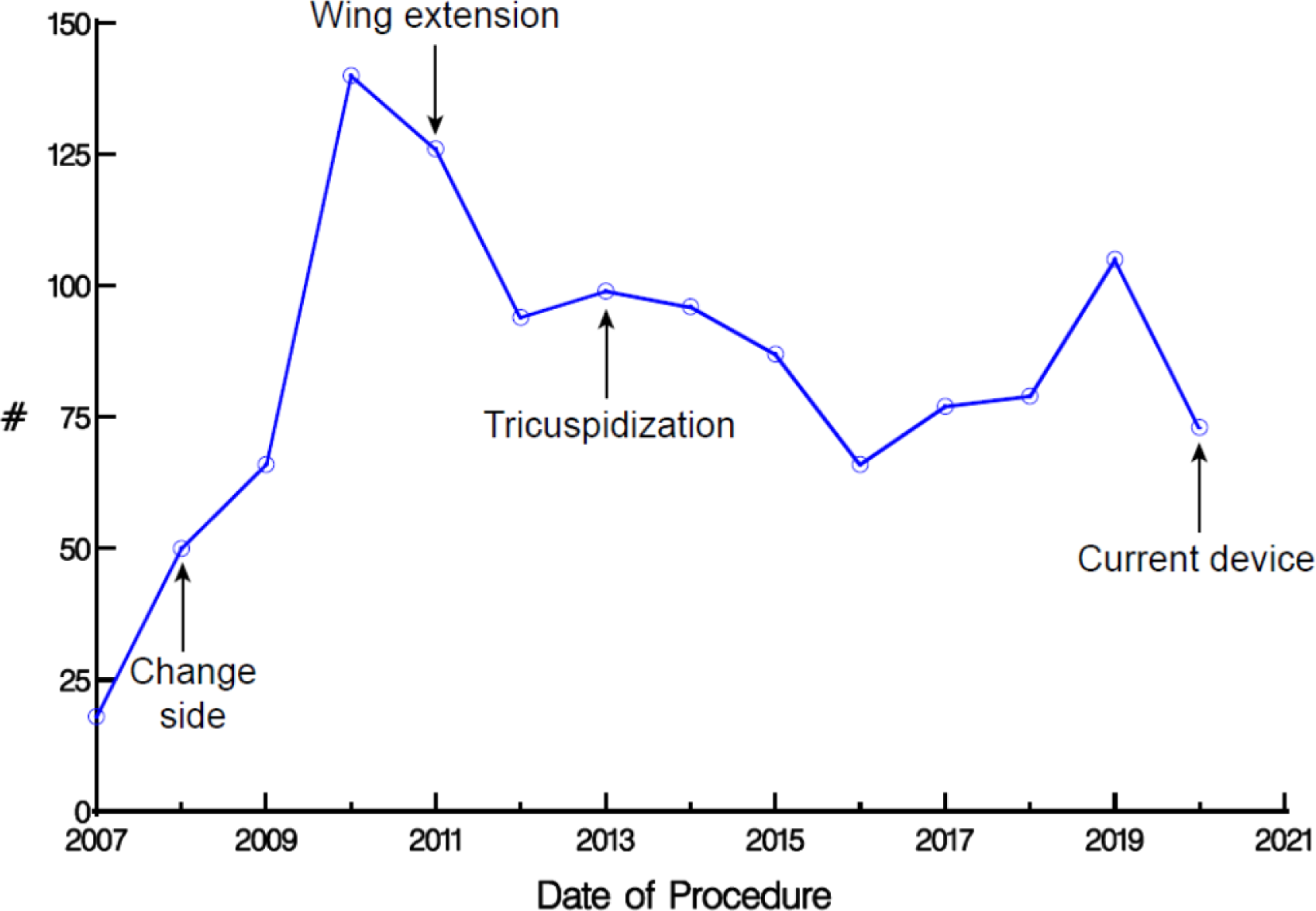
Technique changes over time and number of patients who underwent an Ozaki procedure during the study period. Symbols depict yearly number of cases. Arrows indicate years in which technical modifications were implemented.

**Figure E2:**
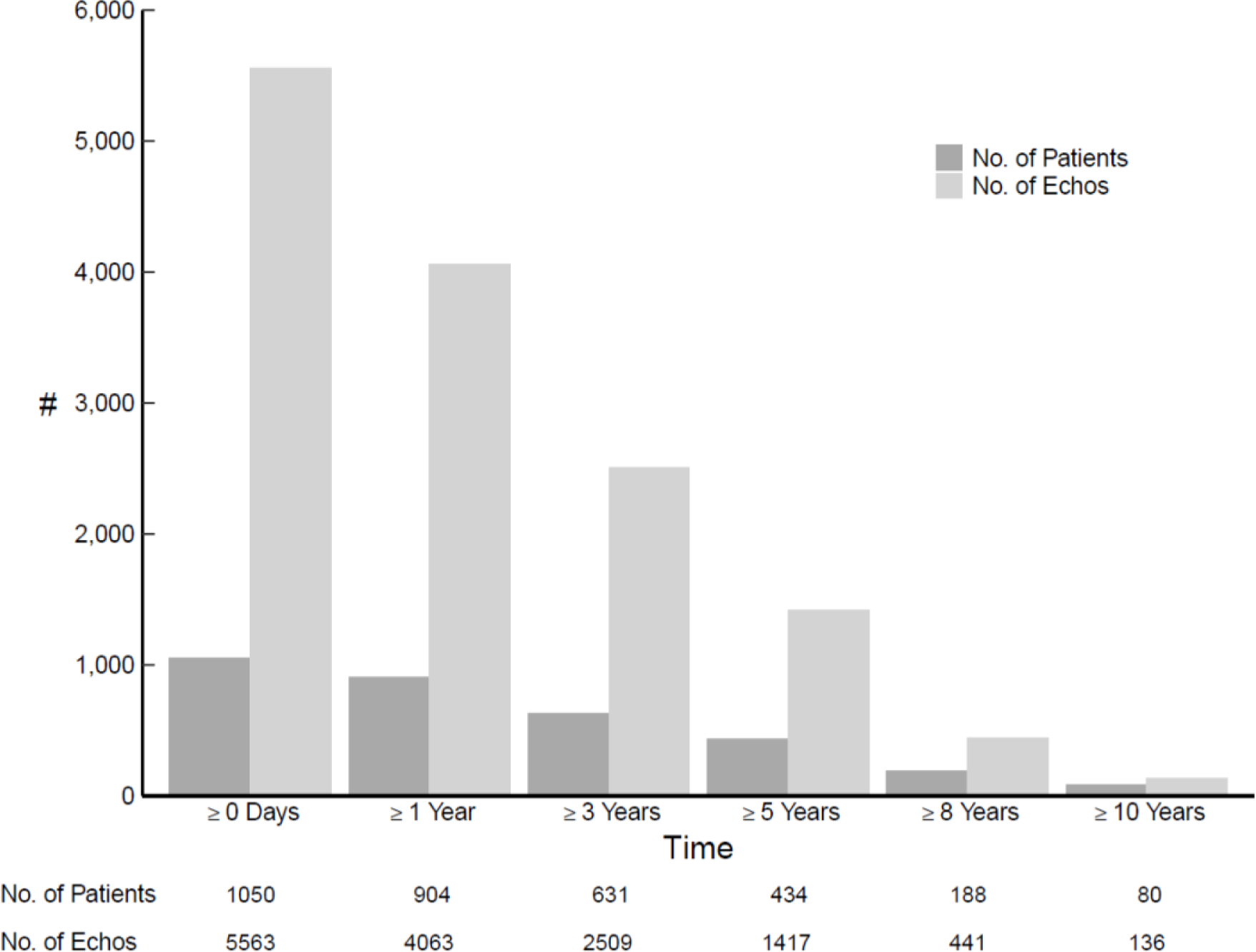
Echocardiography follow-up across study period. Graph shows gradual attrition of patients having echocardiography over time, and number of follow-up echocardiograms (Echos) available at and beyond designated time points.

**Figure E3:**
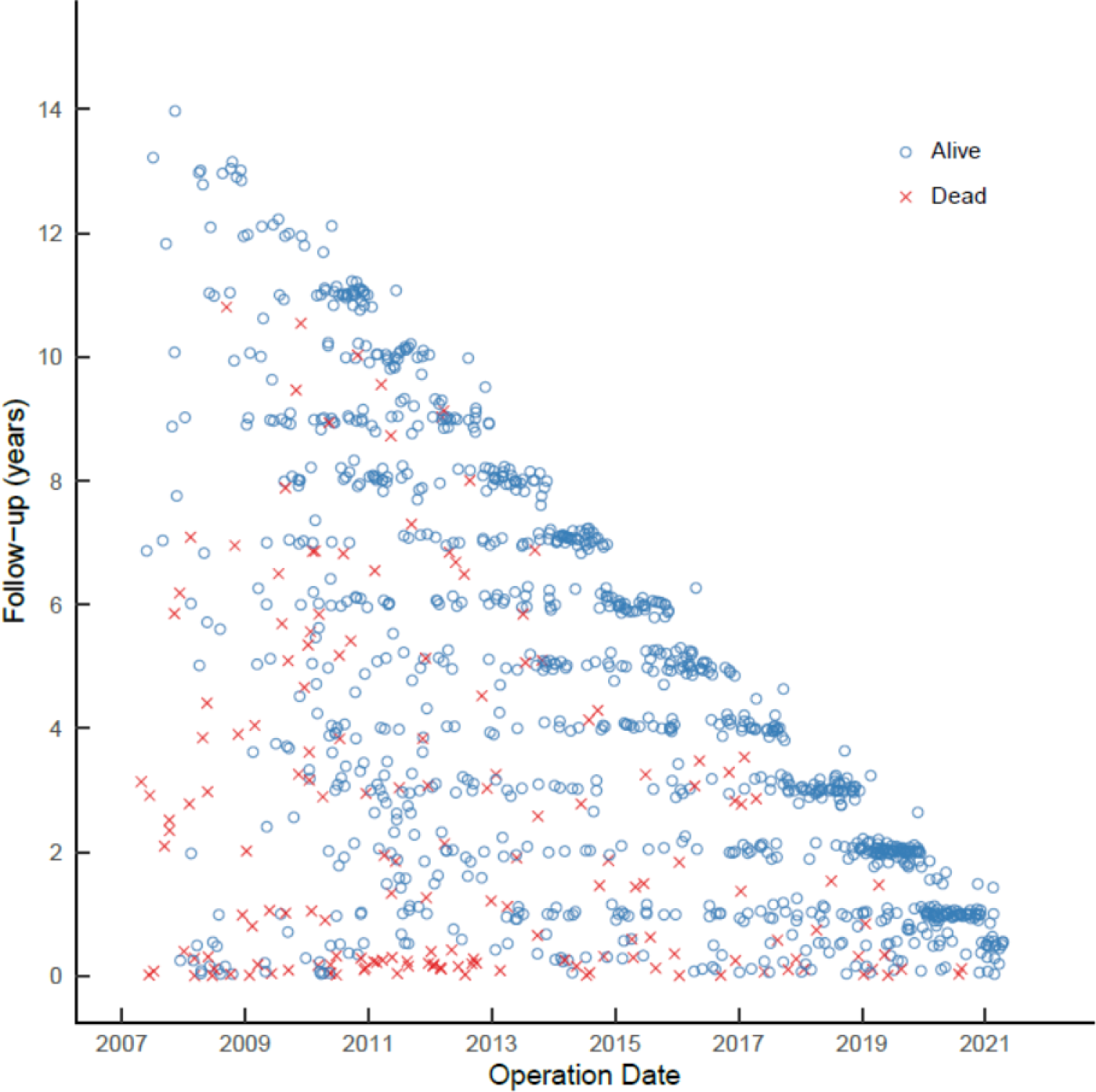
Completeness of follow-up for reoperation and death across study period. Blue circles along diagonal represent compete follow-up, and isolated blue circles are patients with varying degrees of less than complete follow-up.

**Figure E4:**
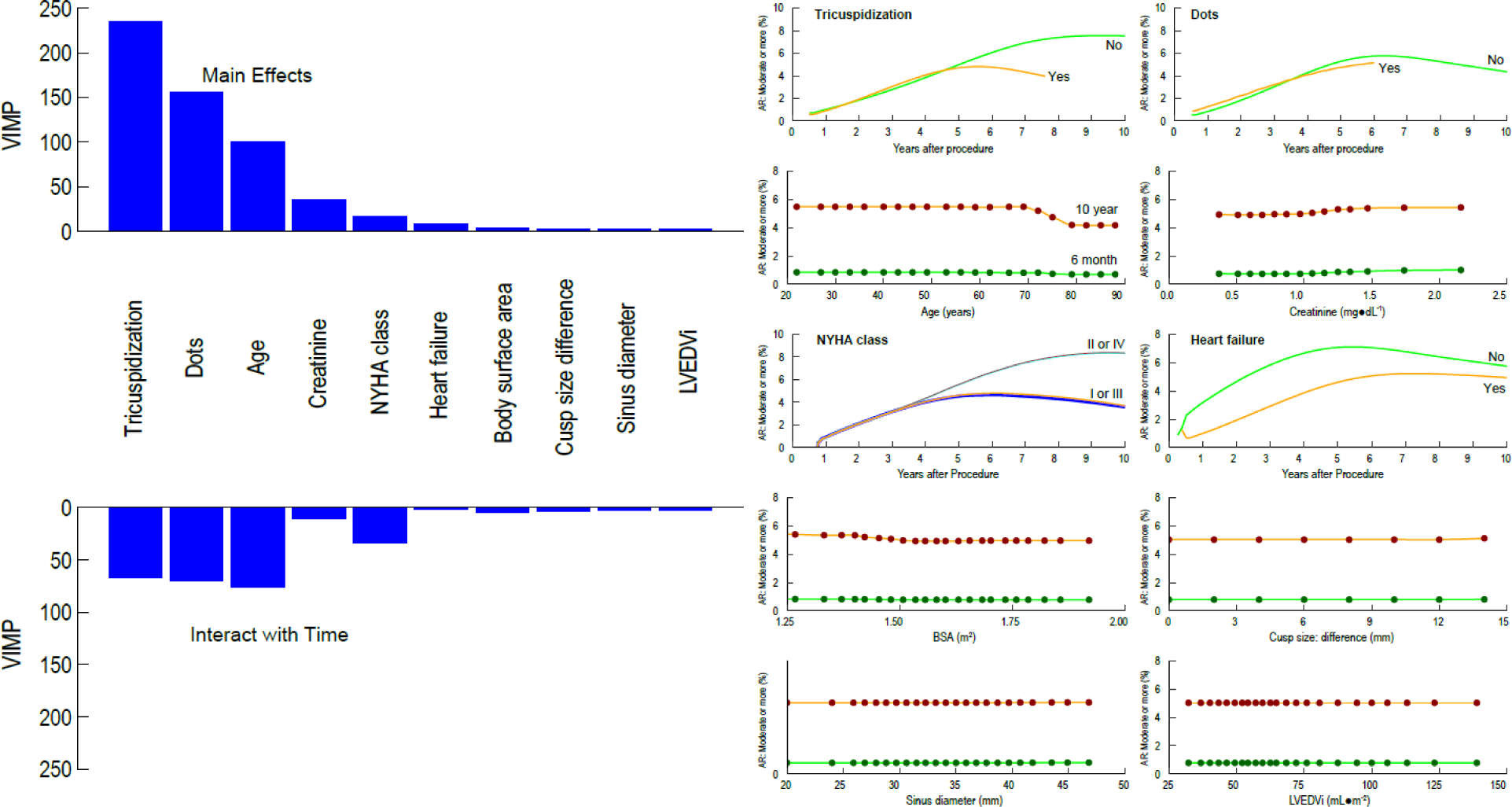
Preoperative and operative variables associated with risk of moderate or greater aortic regurgitation (AR) after Ozaki procedure. Left panel depicts variable importance (VIMP) of the top 10 variables. Variable and bars above the “0” line depict VIMP of variables that may not change with time (main effects); variable and bars below the “0” line depict VIMP of variables whose effect changes with time (interaction with time). Right 2 columns are partial dependency plots depicting the risk-adjusted association between selected patient variables and moderate or greater AR. Symbols are risk-adjusted 6-month (green) and 5-year (brown) predicted probability of moderate or greater AR after Ozaki procedure for different values of selected continuous covariables. Solid lines are smoothed lowess lines of the symbols. Green and orange lines without symbols depict risk-adjusted predicted probability of moderate or greater AR over time for different values of categorical covariables. **Key:** *Dots*, Dot technique modification; *LVEDVi,* left ventricular end-diastolic volume index; *NYHA,* New York Heart Association; *VIMP,* variable importance.

**Figure E5:**
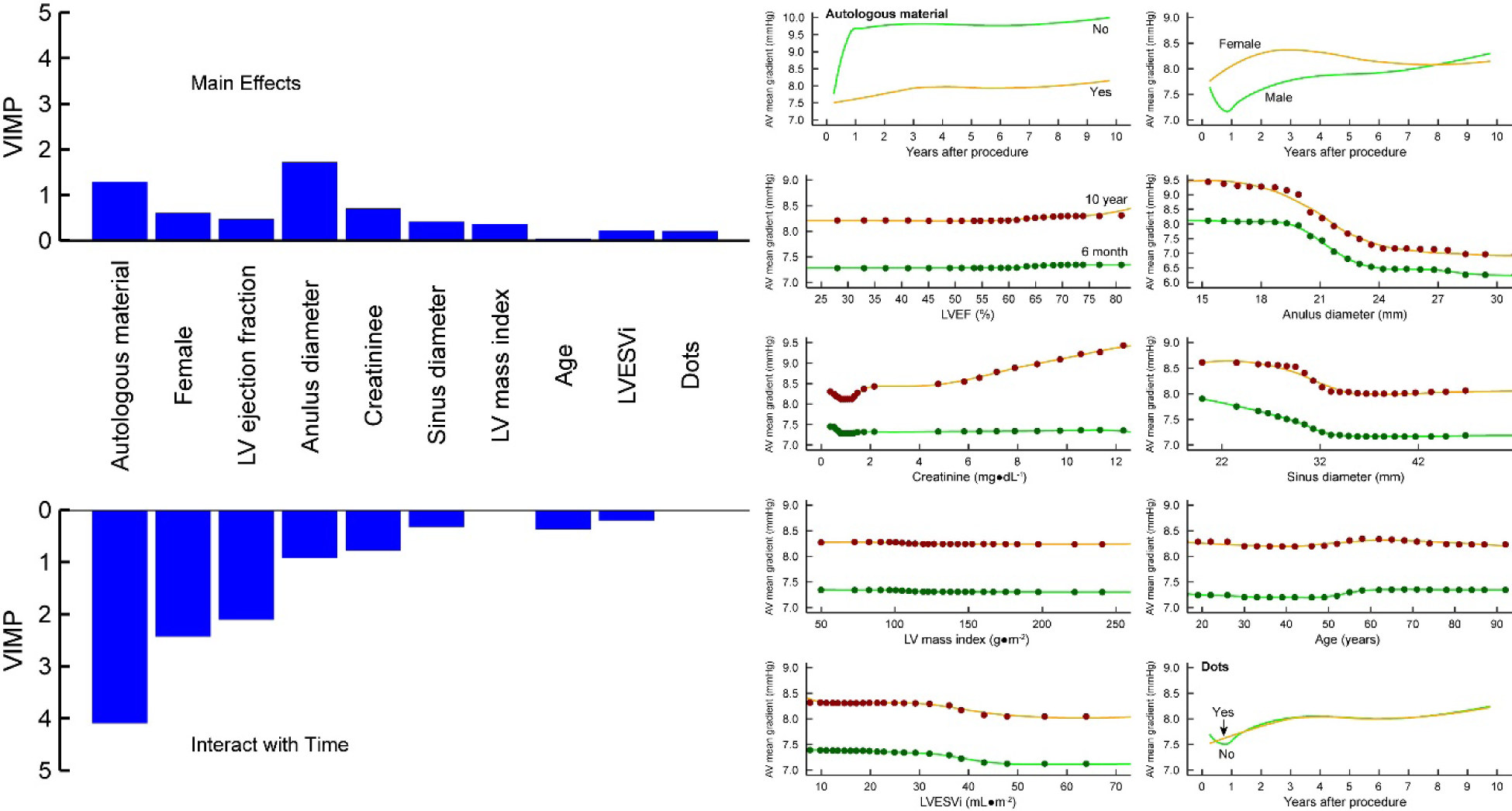
Preoperative and operative variables associated with aortic valve mean gradient after Ozaki procedure. Format as in Figure E4. **Key:** *AR*, aortic regurgitation; Dots, Dot technique modification; *LVEF,* left ventricular ejection fraction; *LV,* left ventricular; *LVESVi,* left ventricular end-systolic volume index; *VIMP*, variable importance.

**Figure E6:**
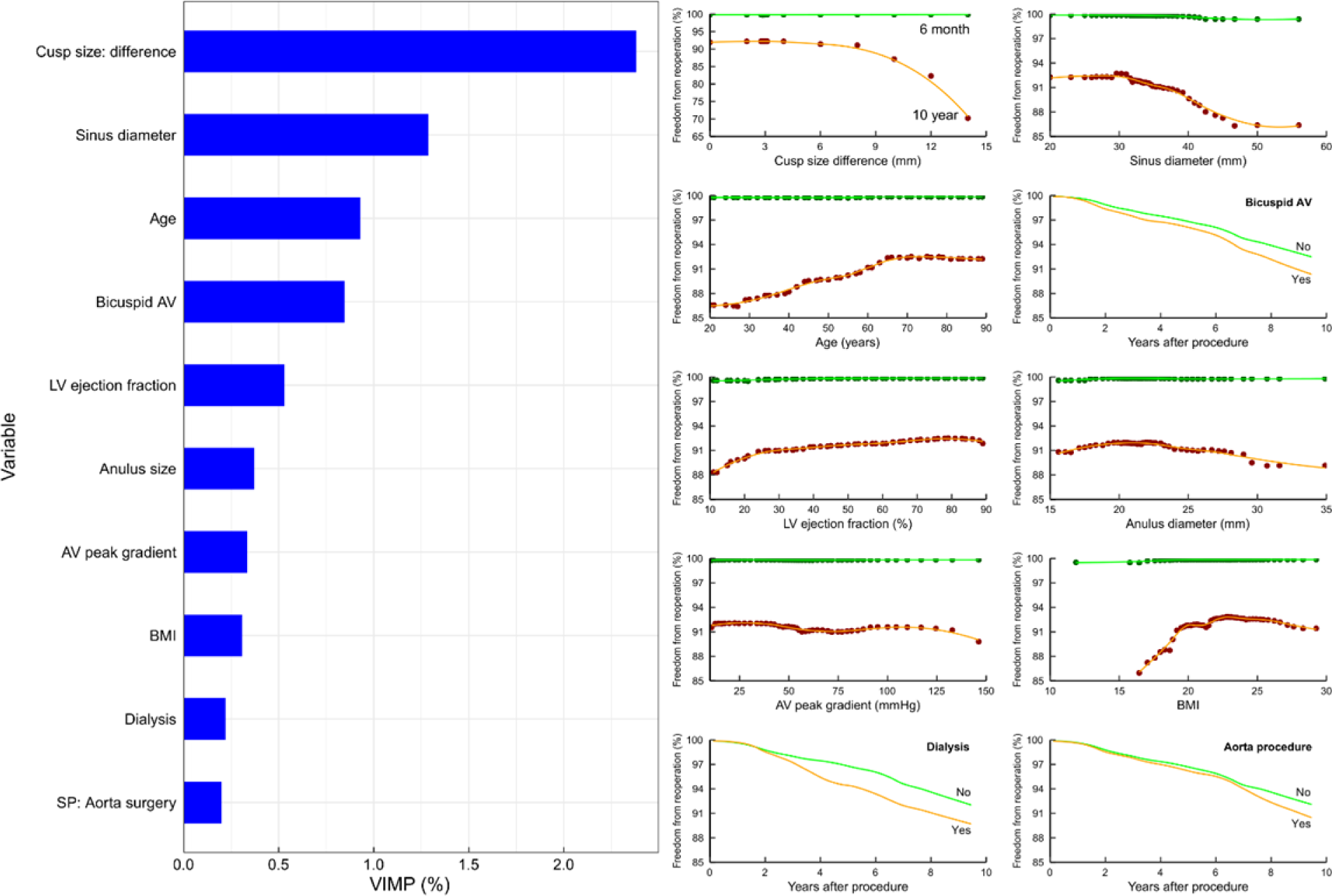
Preoperative and operative variables associated with risk of reoperation. Left panel depicts variable importance (VIMP) of top 10 variables. Right 2 columns are partial dependency plots depicting the risk-adjusted association between selected patient variables and freedom from reoperation. Symbols are risk-adjusted predicted 6-month (green) and 10-year (brown) freedom from reoperation for different values of selected continuous covariables. Solid lines are smoothed lowess lines of the symbols. Green and orange lines without symbols depict risk-adjusted predicted freedom from reoperation over time for different values of categorical covariables. **Key:** *LV,* left ventricular; *VIMP,* variable importance.

**Figure E7:**
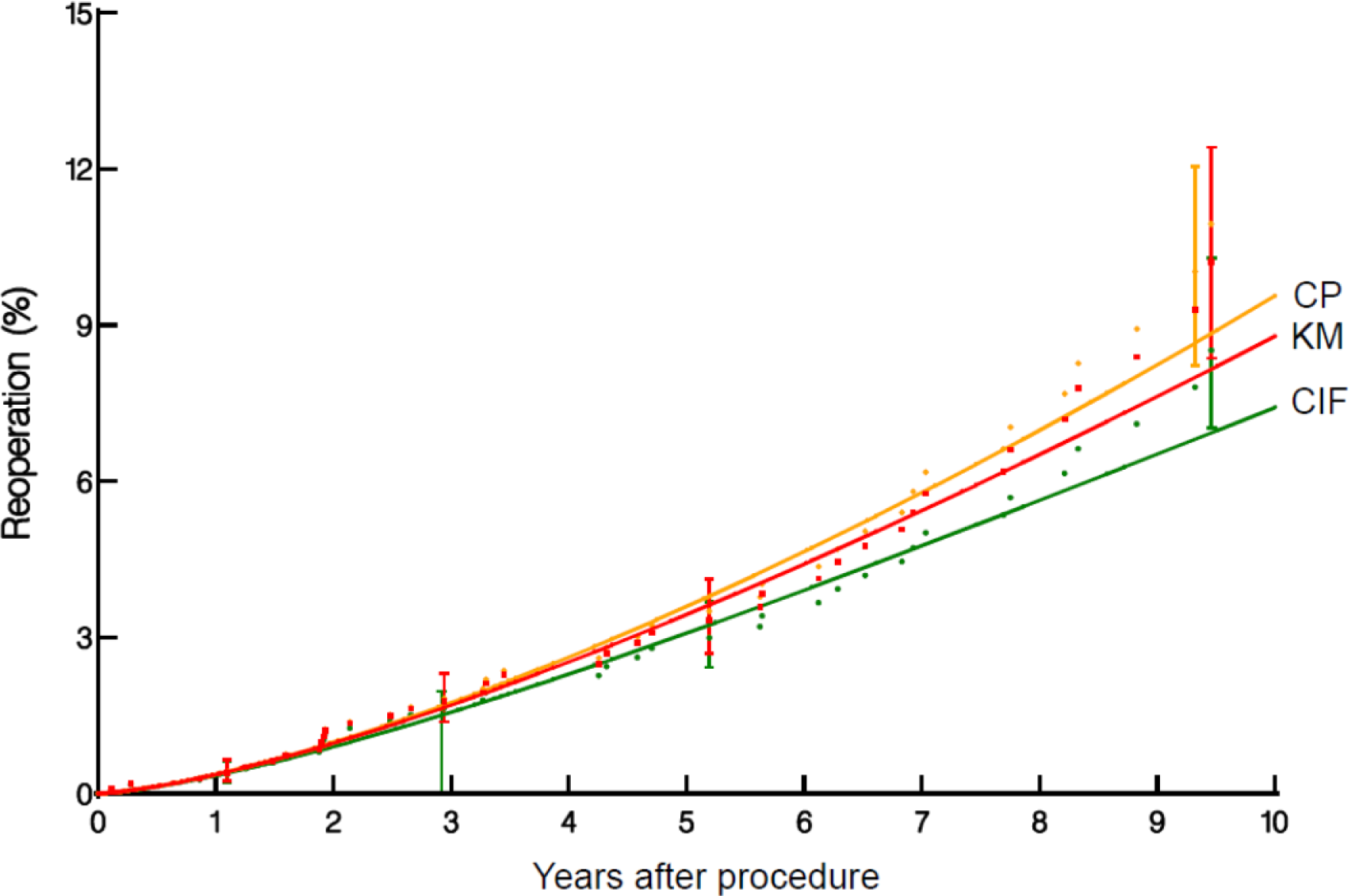
Reoperation after Ozaki procedure. The Kaplan-Meier-type curve **(KM**) (red) depicts the probability of this event, ignoring competing events, thus demonstrating the pure/hypothetical probability of reoperation. The cumulative incidence function curve **(CIF)** (green) depicts the likelihood of patients having reoperation given the competing risk of death before reoperation. The conditional probability curve (**CP**) (yellow) describes the likelihood of reoperation if no competing risks have occurred.

**Figure E8.**
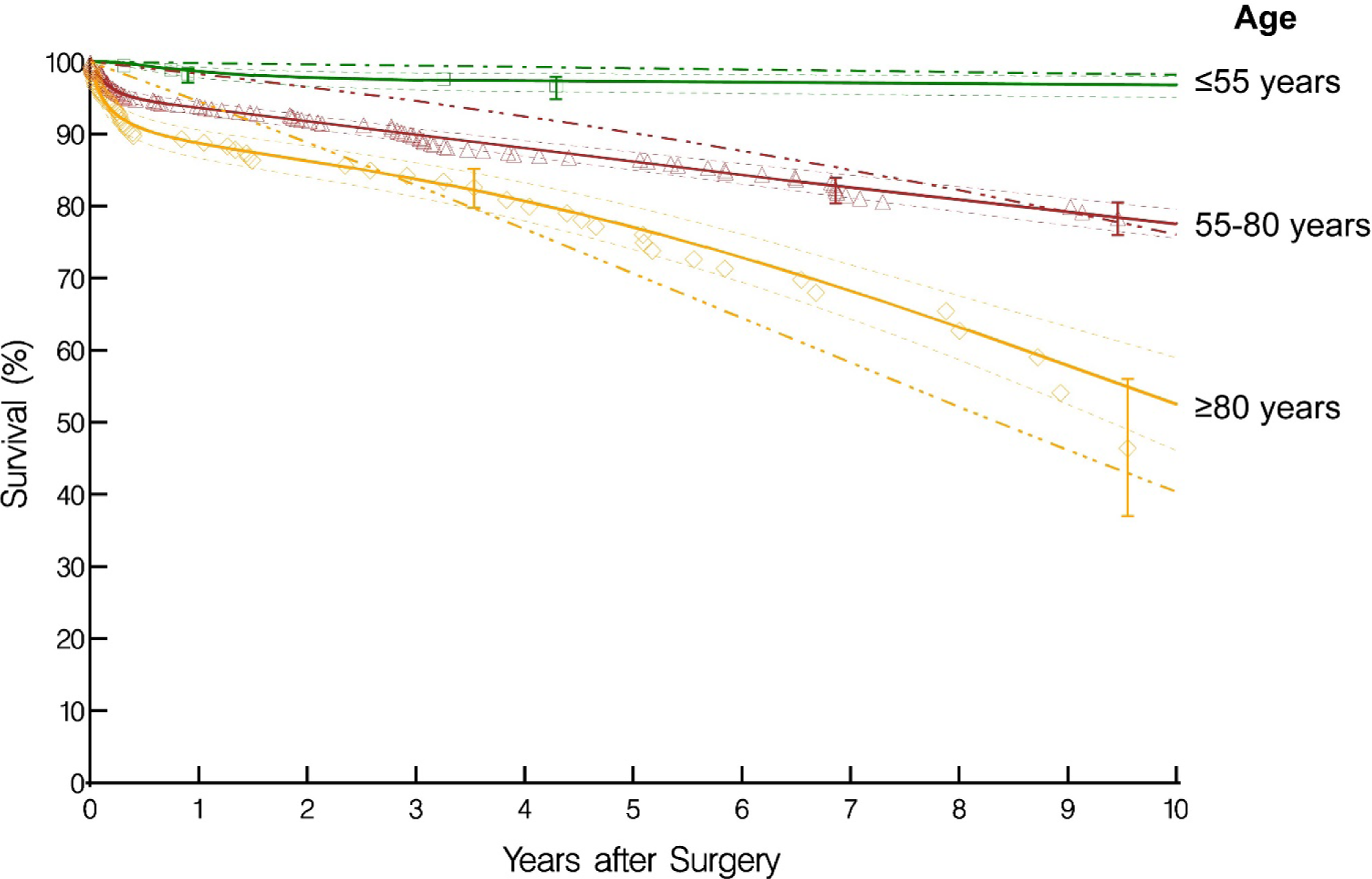
Survival after Ozaki procedure stratified by age groups. Solid lines are parametric estimates enclosed within a 68% confidence band. Kaplan-Meier nonparametric estimates are given by the respective symbols enclosed within the 68% confidence bars equivelent to ±1 standard error at selected time points. Dash-dot-dot line depicts the corresponding expected survival of an age-sex–matched Japanese population, showing that older patients who have undergone an Ozaki procedure have better life expectancy than their counterparts.

**Figure E9.**
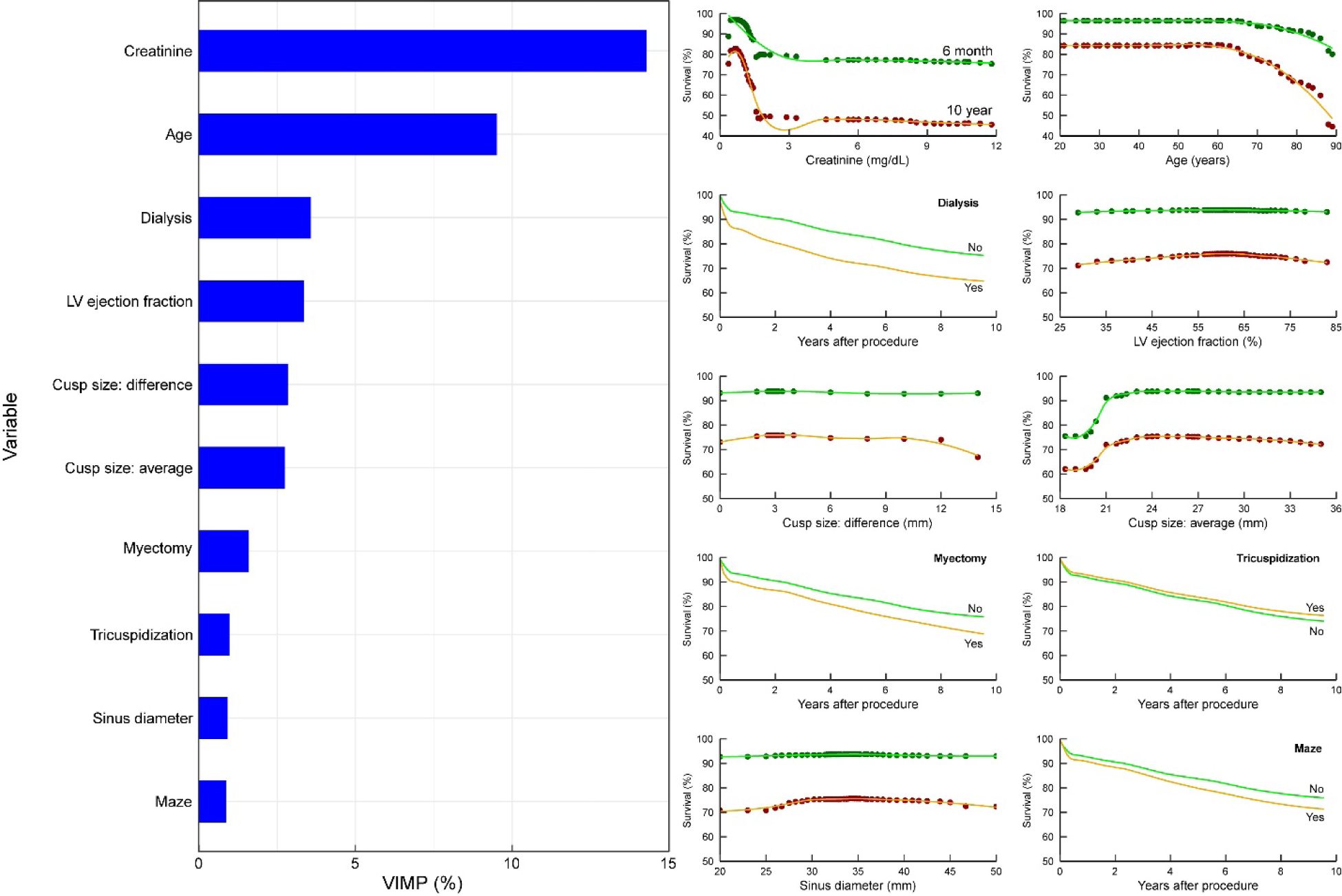
Preoperative and operative variables associated with risk of death. Format as in Figure E6. **Key:** *LV,* left ventricular.

## Appendix E1: Variables Considered in Multivariable Analyses

### Demographics

Female, age (y), body surface area (m^2^), body mass index (kg/m^2^)

### Symptoms

New York Heart Association functional class (I-IV)

### Comorbidities

Previous cardiac operation, previous cardiac interventions (operation or percutaneous coronary intervention), tobacco use (never, current, former), diabetes, dialysis, history of hypertension, heart failure within 2 weeks before surgery, creatinine (mg/dL)

### Aortic valve morphology

Unicuspid, bicuspid, tricuspid

### Aortic valve pathology

Aortic regurgitation grade, pure aortic regurgitation, pure aortic stenosis, mixed aortic regurgitation–aortic stenosis, other

### Echo: Aorta dimensions/Aortic valve hemodynamics

Aortic anulus (mm), sinus of Valsalva (mm), aortic valve peak pressure gradient (mmHg), aortic valve mean pressure gradient (mmHg)

### Cusp size

Size difference, size average

### Echo: Ventricular dimensions/function/mass

Left ventricular (LV) end-diastolic volume–Teichholz (mL), LV end-diastolic volume index (mL/m^2^), LV end-systolic volume–Teichholz (mL), LV end-systolic volume index (mL/m^2^), LV ejection fraction–Teichholz (%), LV mass index (g/m^2^)

### Operative details

Coronary artery bypass grafting, aortic root, aortic procedure, maze, tricuspid valve procedure, mitral valve procedure, atrial septal defect/patent foramen ovale closure, myectomy, technical modification: wing extension, tricuspidization, Dot technique modification

### Pericardium material

Autologous, bovine, equine

